# Retrospective study of COVID-19 seroprevalence among tissue donors at the onset of the outbreak before implementation of strict lockdown measures in France

**DOI:** 10.1101/2020.09.11.20192518

**Authors:** Nicolas Germain, Stéphanie Herwegh, Anne-Sophie Hatzfeld, Laurence Bocket, Brigitte Prévost, Pierre-Marié Danze, Philippe Marchetti

## Abstract

**Background:** The COVID-19 pandemic has altered organ and tissue donations as well as transplantation practices. SARS-CoV-2 serological tests could help in the selection of donors. We assessed COVID-19 seroprevalence in a population of tissue donors, at the onset of the outbreak in France, before systematic screening of donors for SARS-CoV-2 RNA.

**Methods:** 235 tissue donors at the Lille Tissue bank between November 1, 2019 and March 16, 2020 were included. Archived serum samples were tested for SARS-CoV-2 antibodies using two FDA-approved kits.

Results: Most donors were at higher risks for severe COVID-19 illness including age over 65 years (142/235) and/or presence of co-morbidities (141/235). According to the COVID-19 risk assessment of transmission, 183 out of 235 tissue donors presented with a low risk level and 52 donors with an intermediate risk level of donor derived infection. Four out of the 235 (1.7%) tested specimens were positive for anti-SARS-CoV-2 antibodies: 2 donors with anti-N protein IgG and 2 other donors with anti-S protein total Ig. None of them had both type of antibodies. Conclusion: Regarding the seroprevalence among tissue donors, we concluded that the transmission probability to recipient via tissue products was very low at the beginning of the outbreak.

## Introduction

First identified in Wuhan (China), in early January 2020, the new severe acute respiratory syndrome virus 2 (SARS-CoV-2), responsible for the coronavirus disease 2019 (COVID-19), rapidly spread to other countries worldwide causing an unprecedented pandemic ^1^. In France, the first confirmed cases of COVID-19 were identified by the National Reference Center on January 24, 2020, in Bordeaux and Paris in persons who had recently stayed in Wuhan ^2^. These imported cases were followed by the onset of new cases who acquired the infection due to subsequent local transmission in Europe, thus confirming an ongoing COVID-19 outbreak ^3^. The situation evolved rapidly, to limit the spread of the virus, on March 16, 2020, the French government declared a full lockdown of commercial and social activities throughout the territory, which ended on May 11, 2020 (Fig. 1).

**Figure 1.**
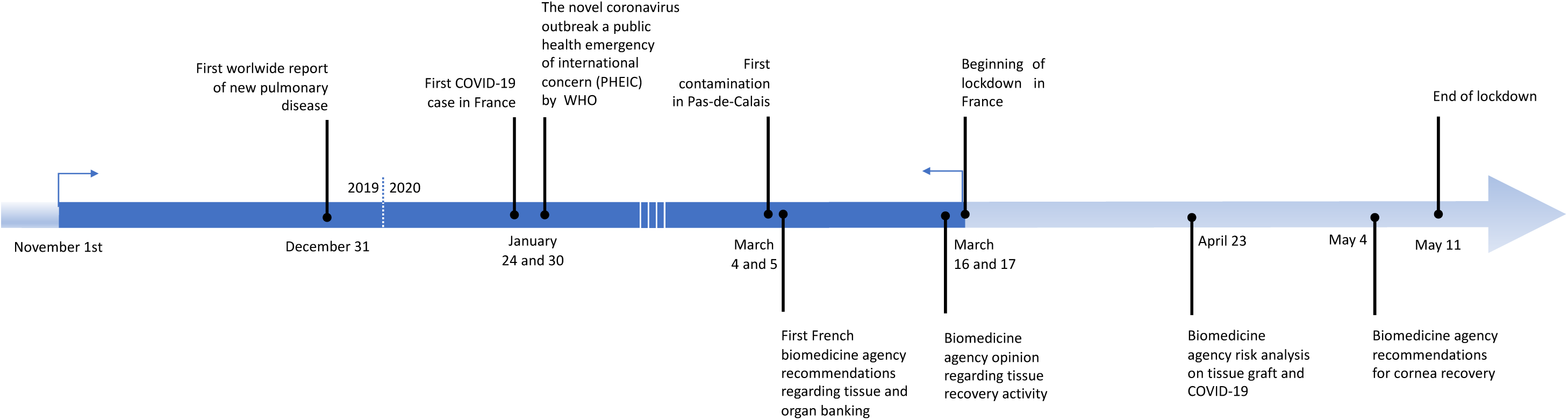
Timeline of the COVID-19 pandemic. Study period in dark blue.

The current COVID-19 pandemic has dramatically modified donation and transplantation practices ^4^. COVID-19 knowledge is constantly evolving and potentially, the SARS-Cov-2 could affect the safety and/or quality of several tissues and organs. SARS-Cov-2 primary infects the lungs and airways. Although the main transmission mode is via person-to-person contact, mainly through respiratory droplets, other transmission modes cannot be excluded. Indeed, SARS-Cov-2 was found in the blood as well as multiple organs and tissues well beyond the respiratory tract ^5^. Thus, in light of these uncertainties, some questions arise concerning the risk of transmission of SARS-CoV-2 through tissue or organ donations. The incertitude about transmission risk via tissue donors is increased by the fact that in most cases, (over 80% of cases) donors are asymptomatic or present with very little symptoms^6^.

Taking into account the information available, the French Biomedicine Agency updated the guidance on SARS-CoV-2 transmission risk via donated organs and tissues on March 5, 2020 and recommended to exclude donors with symptoms suggestive of COVID-19 (fever, cough, etc.) and donors who had stayed or traveled to high risk regions within the prior 28 days, or were in direct contact with known or suspected COVID-19 cases within the prior 28 days. On March 15, updated recommendations called for the systematic detection of SARS-CoV-2 by RT-PCR for all potential donors (Fig. 1).

Since donor testing for COVID-19 was not systematically realized pre-procurement before March 15, 2020, some individuals a priori eligible for tissue donation with mild or asymptomatic COVID-19 could have remained undetected during screening.

While the detection of SARS-CoV-2 nucleic acid by PCR in nasopharyngeal swabs is the reference method for the diagnosis of acute COVID-19 infection, recent data suggest that the identification of anti-SARS-CoV-2 antibodies, now widely available, could be useful in assessing the extent of infection in subpopulations ^7^ Notably, serology tests can help estimate whether donors were previously infected even in the absence of symptoms. Seroconversion to SARS-CoV-2 occurs approximatively 1-2 weeks post symptom onset and the antibodies persist for several months ^8^. Several laboratory tests with different performance characteristics received an Emergency Use Authorization delivered by the U.S. Food and Drug Administration (FDA) and/or CE marking for European countries. These serologic tests differ on the type of antibodies detected and on antigen specificity. To date, there is no clear advantage to use assays test for IgG, IgM and IgG, or total antibody since kinetics of each isotype remain poorly known. A correct interpretation of serologic assays depends on their antigen specificity. Schematically, anti-SARS-CoV-2 antibodies target two main viral proteins: the N protein, an internal nucleocapsid protein of the virus, or the S (spike) proteins, surface proteins that bind host cells (via RBD domains) and mediate virus entry into target cells ^9^. S protein Antibodies are more specific than N protein ones, their kinetics also differ. For these reasons, it is advisable to use two tests, one targeting the S protein and one targeting the N protein. According to recently published CDC guidelines ^10^, it is also recommended to use tests with a specificity over 99.5%, which ensures a high predictive value of a positive test in populations low COVID-19 prevalence.

We performed a retrospective cohort study of tissue donors at the Tissue Bank of the university hospital of Lille, France to assess COVID-19 risk. We estimated the seroprevalence of antibodies to SARS-CoV-2 in tissue donors before the systematic implementation of RT-PCR SARS-CoV-2 in all donors. This study documents the risk assessment of tissue donation in a period of community transmission and helped determine the fate of stored tissues.

## Materials and methods

### Donors and data collection

We reviewed the medical files of 235 tissue donors processed at the Lille tissue bank between November 1, 2019 and March 16, 2020 (Fig.1 and 2). Donations were distributed over the months as such: 47 in November, 44 in December, 52 in January, 56 in February and 36 in March. The following data were collected and analyzed: type of donation, age, gender, place of death, cause of death, medical history to assess COVID-19 risk, virology testing, number and type of specimen recovered.

**Figure 2.**
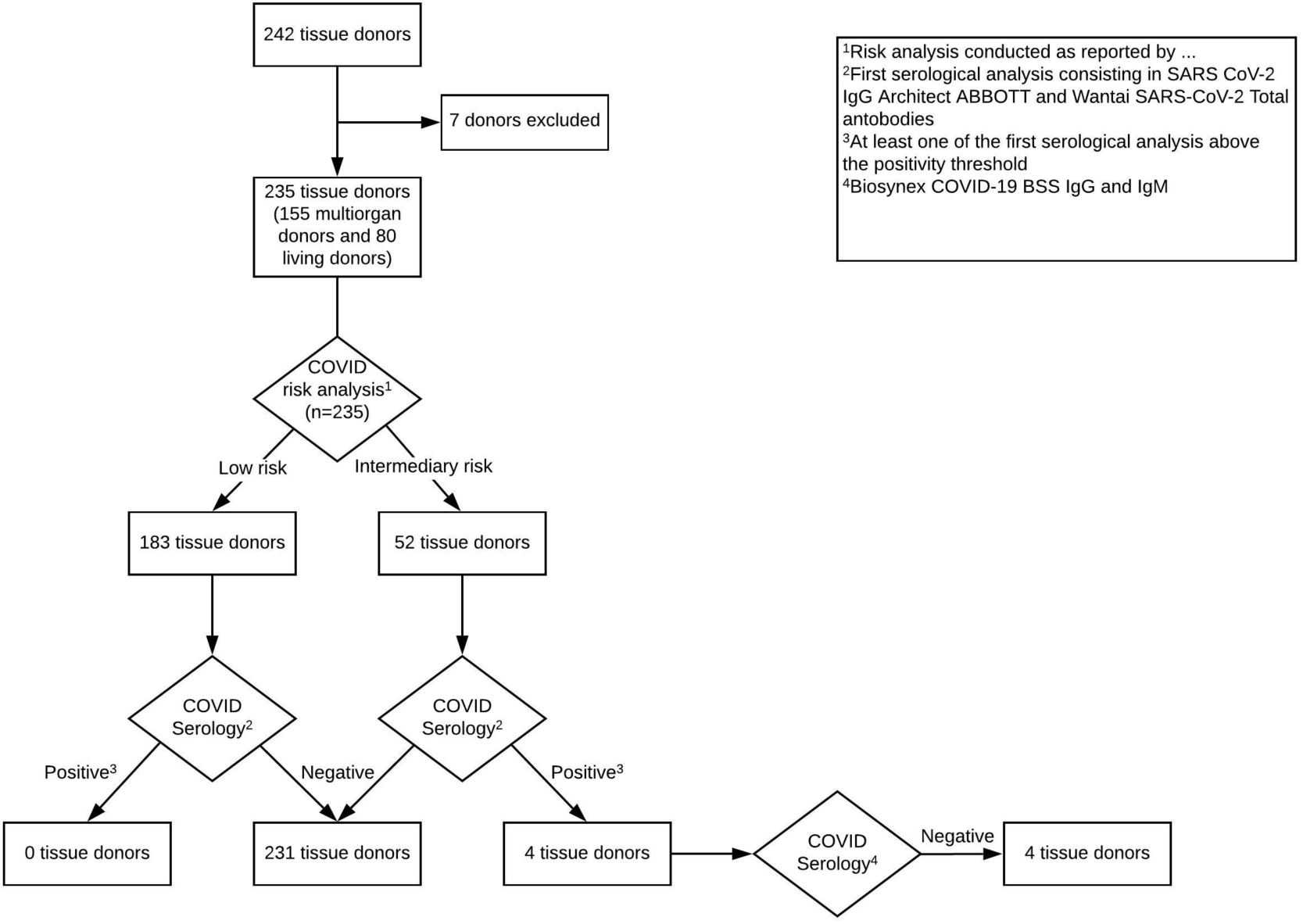
Flowchart of risk analysis and serological testing. 242 tissue donors were studied between November 1, 2019 and March 16, 2020. Seven donors were excluded for insufficient blood collection. 235 tissue donors were divided into low (n=183) and intermediary (n=52) COVID-19 risk and tested for COVID-19 antibodies. Overall, 231 tissue donors were negative and 4 had a positive serology.

Epidemiological characteristics of COVID-19 infection in “Hauts de France” (location of first cases, number of cases at lockdown, evolution of cases in “Nord” and “Pas De Calais”) were also collected^11^.

### Analysis of anti-SARS-CoV-2 antibodies

Archived blood specimens collected on the day of donation for donor screening of infectious diseases were retrospectively tested for SARS-CoV-2 antibodies (Fig.2). Anonymized archived specimens were stored at -80°C until the SARS-CoV-2 antibody testing was performed. Archived samples from 235 donors (7 donors were excluded because of insufficient serum volumes) were screened for the presence of anti-SARS-CoV-2 by two complementary FDA-approved immunoassays with a specificity >99.5 %. The SARS-CoV-2 chemiluminescent microparticle immunoassay (CMIA) (Batch #17298FN00, controls batch #17100FN00 and #17531FN00, Abbott, Illinois, USA) is designed to detect immunoglobulin class G (IgG) antibodies to the nucleocapsid (N) protein of SARS-CoV-2. The presence or absence of IgG antibodies to SARS-CoV-2 in the sample is determined by comparing the chemiluminescent relative light unit (RLU) in the reaction to the calibrator RLU. Results are interpreted as positive (ratio ≥ 1.4) or negative (ratio < 1.4). External positive controls and donor samples were analyzed on an Abbott Architect i2000SR according to the manufacturer’s instructions.

Performance characteristics of this test are 100% sensitivity and 99.6% specificity according to the FDA ^12^. The SARS-CoV-2 total Ab Elisa test (Batch #NCOA20200301 and #NCOM20200301, Beijing Wantai Biological Pharmacy Enterprise, Beijing, China; Cat # WS1096) was also used. The Wantai SARS-CoV-2 Ab is an enzyme-linked immunosorbent assays (ELISA) in a 96-well-plate format detecting total antibodies (including IgM and IgG) binding SARS-CoV-2 spike protein receptor binding domain (RBD). Results are calculated by relating each specimen absorbance (A) value to the Cut-off value (C.O.) of the plate. C.O. is calculated by mean absorbance value for three negative controls plus 0.16. The presence or absence of IgG antibodies to SARS-CoV-2 in the sample is determined by comparing the absorbance in the sample to C.O. values. Results are interpreted as positive (ratio ≥ 1) or negative (ratio < 1). Specimens with absorbance to Cut-off ratio between 0.9 and 1.1 are considered borderline and duplicate retesting of these specimens is required to validate initial results. External positive controls and donor samples were prepared and analyzed on an Quadriga BeFree and BEPIII (Siemens, Munich, Germany) according to the manufacturer’s instructions. The Wantai Total Ab ELISA performance is 94.5% sensitivity and 100% specificity, with PPV:100% and NPV 95.33% as reported by the manufacturer.

All tests were performed using the same lot. No detection for SARS-CoV-2 was done through direct RT-PCR on nasopharyngeal and oropharyngeal swabs.

### Ethics statement

This study was performed according to the Declaration of Helsinki and approved by the Medical Ethical Committee of the CHU de Lille. Written informed consent forms were obtained from all enrolled living donors.

## Results

### Donor characteristics

Our retrospective study was carried out at the Lille Tissue Bank on the entire group of tissue donors over a period ranging from November 1, 2019 to March 16, 2020, prior to the lockdown (Fig. 1). Figure 3 shows the geographic location of donors throughout the Hauts-de-France region. None of the donors came from COVID-19 hot spots, most of them came from areas where only few cases of COVID-19 were reported as of March, 5, 2020. Table 1 summarizes the main characteristics of the 235 donors included in this study. Most donors were women (122 [51. 9 %]). Median age of donors was 68 years (IQR 79-57). Among the 156 deceased donors, sepsis was the principal cause of death (19.9 %) followed by stroke (17.3%), respiratory failure (14.7 %) and heart attack (12.8 %). Next, we screened the donors for underlying conditions important to the severity of COVID-19 ^6^. Most donors were at higher risk for severe COVID-19 illness. In all, 142 (60.4 %) of the donors were aged 65 years and over. Among 141 donors with underlying conditions, 70 presented with high blood pressure (46.4 %), 39 with diabetes (25.8 %) and 6 with obesity (4 %). Cardiovascular disease was found in 54 donors (35.8 %), chronic lung disease in 37 donors (24.5 %) and malignant cancer in 25 donors (16.6 %). Furthermore, 16 donors (10.6 %) presented with chronic kidney disease or liver disease.

**Figure 3.**
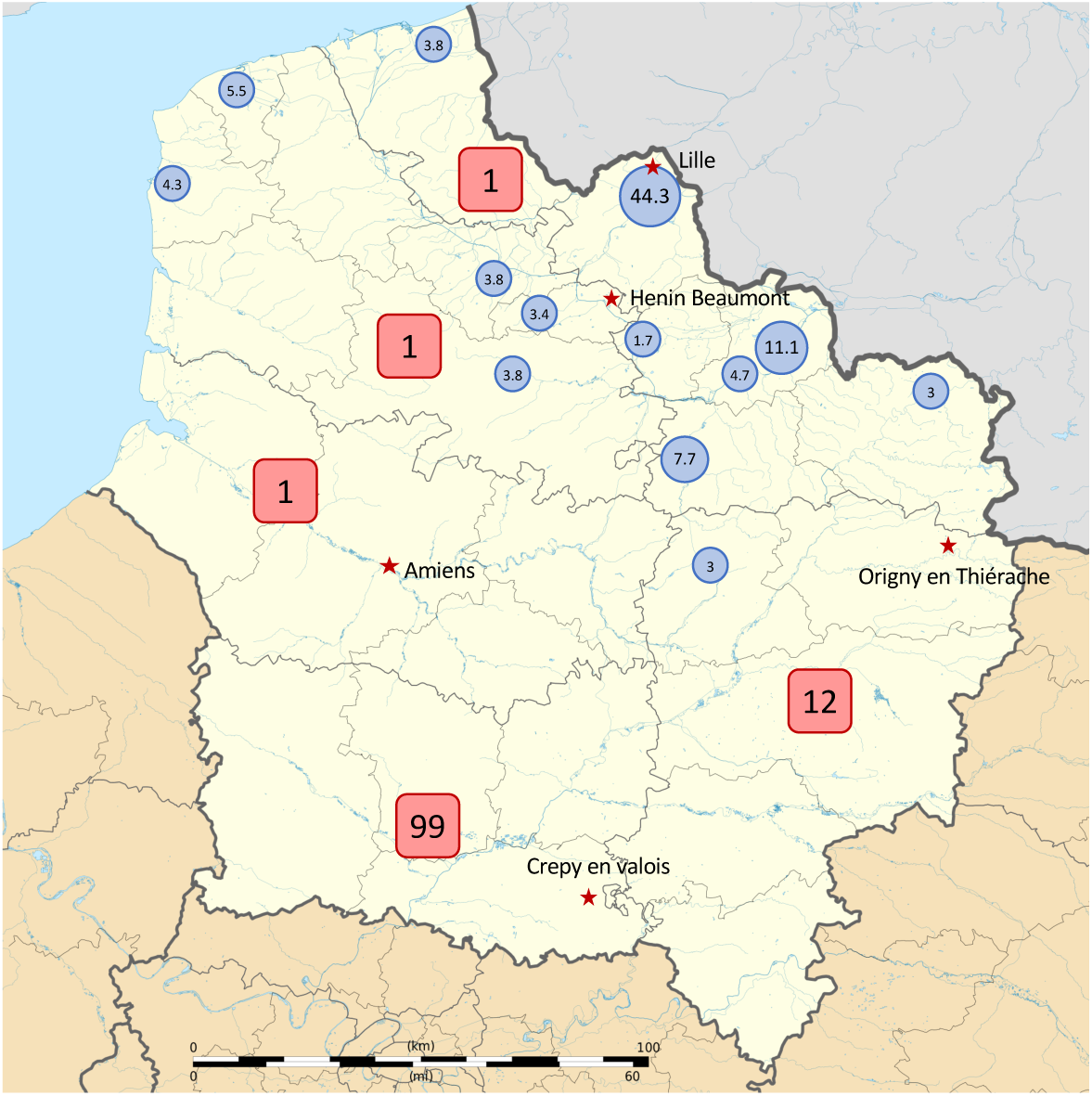
Map of COVID cases and tissue donors in « Haut de France » region. Cumulative number of COVID-19 confirmed cases on March 5, 2020 in each department (Aisne, Nord, Oise, Pas-de-Calais, Somme) framed in red. First cases of COVID-19 in each department are represented by a red star. Proportion of the 325 studied tissue donors regarding location of recovery in blue circles.

**Table 1.**
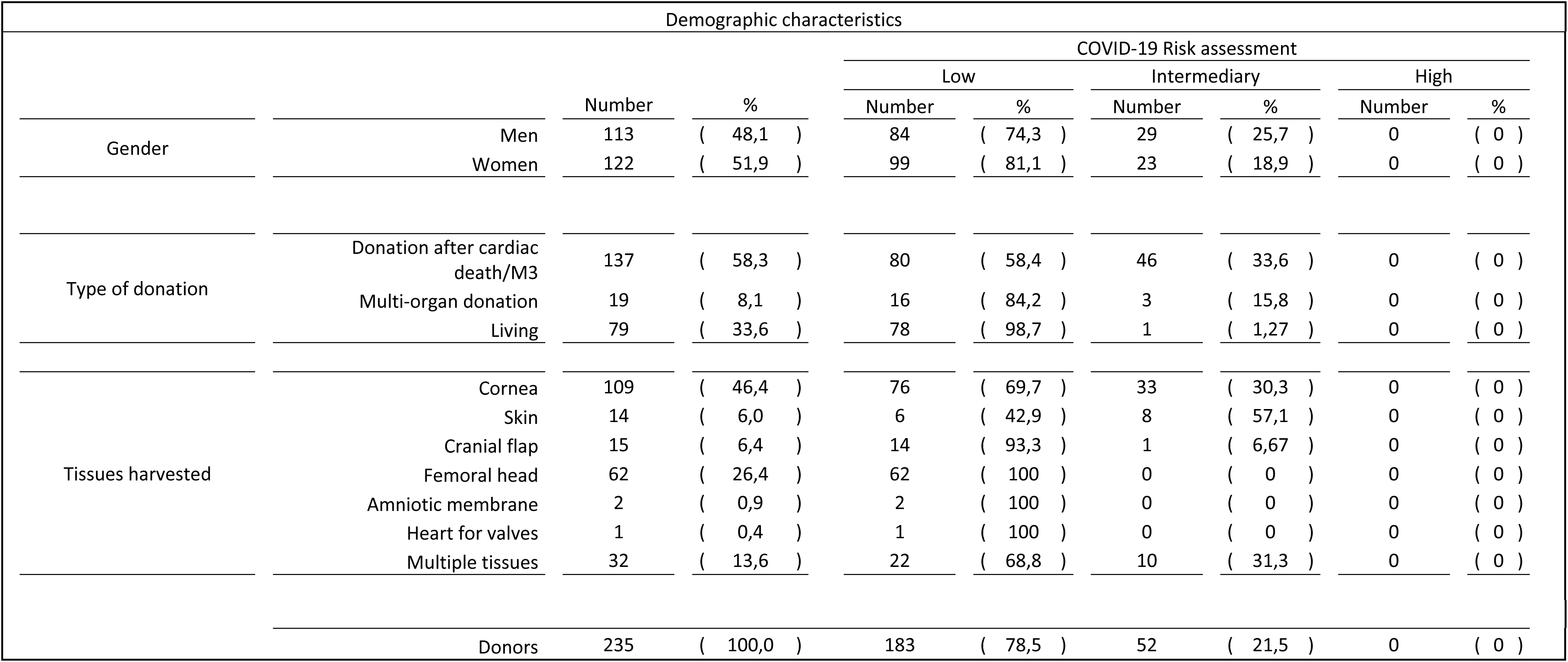
Demographic characteristics of 235 tissue donors

### Characteristics of donors with intermediate risk of COVID-19 infection

Next, we assessed the COVID-19 risk profile in the donor population. According to the American Society of Transplantation ^13^, we retrospectively used clinical and epidemiological criteria to stratify donors into high, intermediate, or low COVID-19 risk of donor derived infection. No donor was classified at high COVID-19 risk. In contrast, we identified 52 donors (21.5 %) at intermediate COVID-19 risk of donor derived infection since they developed COVID-19 compatible symptoms (Table 1). Most common signs were acute respiratory distress (23 donors), dyspnea (14 donors), other flu-like/respiratory symptoms (18 donors), chest CT image opacity without clinical symptoms (2 donors), cough (1 donor), fever (1 donor). Other tissue donors (183/235) were considered at low COVID-19 risk.

### Presence of anti-SARS-CoV-2 antibodies in serum samples of tissue donors

Archived serum samples were used to test retrospectively the new infectious agent, SARS-CoV-2. Serum samples of the 235 donors collected on the day of donation were tested for antibodies against SARS-CoV-2. Only 4 out of 235 (1.7 %) samples contained anti-SARS-CoV-2 antibodies: 2 samples presented with IgG directed against the full length nucleocapsid protein and 2 other samples were weakly positive for anti-SARS-CoV-2 total Ig directed against the S1 domain of viral spike protein (Table 2). Screening serology (including testing against hepatitis B and C, syphilis, HIV-1 and -2, and HTLV) was negative excluding potentially cross-reacting antibodies. Out of the 4 seropositive donors, two had an intermediate COVID-19 risk since they developed COVID-19 -related symptoms such as fever or acute respiratory distress syndrome (Table 2).

**Table 2.** Characteristics of the 4 SARS-COV2 serology positive tissue donors

| Positive serology donors characteristics |  |  |  |  |  |  |  |
| --- | --- | --- | --- | --- | --- | --- | --- |
| Gender | Age | Type of donation | COVID risk assessment | Date of serological testing (mm/dd/yy) | Days between symptoms onset and serology | Abott Architect IgG<sup>1</sup> | Wantai total antibodies<sup>2</sup> |
| F | 56 | Multi-organ donation | Low | 11/23/19 | NA | 3.79 | Negative |
| M | 35 | Living | Intermediary | 11/24/19 | Unknown | Negative | 1.84 |
| M | 59 | Living | Low | 12/09/19 | NA | 1.73 | Negative |
| F | 61 | Donation after cardiac death | Intermediary | 03/09/20 | 9 | Negative | 1.9 |
| <sup>1</sup> | Abott Architect IgG positivity threshold $\geq 1.4$ | | | | | | |
| <sup>2</sup> | Wantai total antibodies positivity threshold $\geq 1.1$ | | | | | | |
| NA | Not Applicable |  |  |  |  |  |  |

## Discussion

To our knowledge, this is the first investigation on SARS-CoV-2 seroprevalence in tissue donors. In our study, many donors were at increased risk of COVID-19 infection (Table 1). However, only few donors tested (4/235) were found to be positive for the SARS-Cov-2 antibodies. According to previous studies, seroprevalence rates of SARS-CoV-2 vary considerably, ranging from 0 to more than 25 %, depending on the population studied and methods used ^14,15^. The low prevalence rate among tissue donors can be explained by the fact that patients with an uncontrolled active infection at the time of donation are not eligible for donation according to our selection protocol reducing the risk of symptomatic donors with active COVID-19 enrolled in this study. Another explanation could be that some donors were tested too close to the onset of symptoms before developing antibodies. Classically, antibodies against SARS-Cov-2 seem to appear on day 7 to day10 after illness onset ^8^ Moreover, it is highly likely that most positive cases from our testing procedure were false positive results for several reasons. Although we used commercially available and FDA-approved tests with a high performance as recommended ^12^, due to the different specificities, results should be interpreted with caution ^16^. For example, the Abbott anti-N Ig G test we used, with a positive predictive value (PPV) = estimated at 92.9% assuming a prevalence of 5% ^12^ would likely identify some false positives but no false negative results (NPV 100%). Obviously, we did not know the prevalence of SARS-CoV-2 antibodies in our population during the study period but the real COVID-19 prevalence was probably less than 5 %. According to official data, in France, on March 15, 2020 only 6378 cumulative positive cases of COVID-19 were detected by RT-PCR out of a population of more than 66 million inhabitants ^17^ In fact, since the positive predictive value is correlated to the prevalence level it is therefore possible that the test identified many false-positive individuals. Thus, results of a single test may not be accurate enough to validate the presence of SARS-CoV-2 antibodies. FDA experts recommend performing a second test, screening for the presence of antibodies targeting a different viral protein, to increase the accuracy of antibody detection. Among the four seropositive donors, results of these two tests were discordant, one positive and the other negative, and none of the positive donors had simultaneously both anti-Nucleocapsid protein IgG and anti-Spike protein total antibodies. These discordant results between both tests used could indicate the existence of false positive results. Alternatively, it could be explained by different antibody kinetics targeting Nucleocapsid- or the Spike-proteins. Antibodies directed against the S protein are produced in more advanced stage of SARS-CoV-2 infection and decrease later than those against the N-protein ^8^.

Due to the important risk of false positivity, positive SARS CoV-2 antibody test results should be validated by other relevant elements such as clinical history. Among the four seropositive donors, two of them had a history of symptoms compatibles with COVID-19. Three out of the four positive donors were tested before the outbreak was declared, at a time when the virus was probably not circulating in Europe suggesting that these results could be false positives. Finally, only one donor (woman aged 61) who was tested at the beginning of the outbreak in March 2020 and presented with clinical symptoms of SARS more than a week before the presence of SARS-CoV-2 antibodies could have had a medical history compatible with the COVID-19 infection. Regardless of these considerations, our study revealed that the risk of donors having been infected by SARS-CoV-2 was very weak. Regarding the prevalence of SARS-CoV-2 antibodies among tissue donors unveiled our study, the probability of transmission to recipients via tissue products is low.

To date, little is known about a potential COVID-19 transmission via tissue donation and the risk of donor-to-recipient transmission of SARS-CoV-2 cannot be totally ruled out. SARS-CoV-2 was isolated from blood samples in up to 15% of cases and therefore, all vascularized tissues could potentially be infected ^6^. Moreover, the angiotensin-converting enzyme 2, the cellular receptor for SARS-CoV-2, is widely distributed among human tissues ^18^. In contrast, although the ocular surface was suggested as a site of SARS-CoV-2 infection, its RNA was not found in the corneal tissues of five COVID-19 patients ^19^. Based on data from other SARS viruses, viral RNA was detected in multiple organs from patients who died of severe SARS-CoV infection ^20^. However, the presence of RNA does not necessarily mean that the virus is active and infectious in all tissues. Indeed, the virus only successfully replicated in the lung and small bowel tissues ^20^. Data on transplant recipients with COVID-19 are scarce. There is one report where liver transplantation from an infected individual without COVID-19 symptoms did not result in disease transmission ^21^. No cases of SARS-CoV-2 transmission via tissue transplants were reported so far. Thus, the critical review of available data on COVID-19 suggests that the transmission risk is low after tissue or organ donation ^22^.

In conclusion, our study aligns with the data from the literature and suggests that tissues collected at the beginning of the outbreak, before lockdown, can be used for grafts, taking into account the individual benefit-risk ratio.

## Data Availability

The datasets generated during and/or analysed during the current study are available from the corresponding author on reasonable request.

## Acknowledgments

This study was supported by the Centre de Biologie Pathologie - CHU de Lille.

